# Causal Associations of Sleep Apnea with Alzheimer’s Disease and Cardiovascular Disease: a Bidirectional Mendelian Randomization Analysis

**DOI:** 10.1101/2023.11.20.23298793

**Authors:** Clémence Cavaillès, Shea J. Andrews, Yue Leng, Aadrita Chatterjee, Iyas Daghlas, Kristine Yaffe

## Abstract

**Background:** Sleep apnea (SA) has been linked to an increased risk of dementia in numerous observational studies; whether this is driven by neurodegenerative, vascular or other mechanisms is not clear. We sought to examine the bidirectional causal relationships between SA, Alzheimer’s disease (AD), coronary artery disease (CAD), and ischemic stroke using Mendelian randomization (MR).

**Methods:** Using summary statistics from four recent, large genome-wide association studies of SA (n=523,366), AD (n=64,437), CAD (n=1,165,690), and stroke (n=1,308,460), we conducted bidirectional two-sample MR analyses. Our primary analytic method was fixed-effects inverse variance weighted MR; diagnostics tests and sensitivity analyses were conducted to verify the robustness of the results.

**Results:** We identified a significant causal effect of SA on the risk of CAD (odds ratio (OR_IVW_) =1.35 per log-odds increase in SA liability, 95% confidence interval (CI) =1.25-1.47) and stroke (OR_IVW_=1.13, 95% CI =1.01-1.25). These associations were somewhat attenuated after excluding single-nucleotide polymorphisms associated with body mass index (BMI) (OR_IVW_=1.26, 95% CI =1.15-1.39 for CAD risk; OR_IVW_=1.08, 95% CI =0.96-1.22 for stroke risk). SA was not causally associated with a higher risk of AD (OR_IVW_=1.14, 95% CI =0.91-1.43). We did not find causal effects of AD, CAD, or stroke on risk of SA.

**Conclusions:** These results suggest that SA increased the risk of CAD, and the identified causal association with stroke risk may be confounded by BMI. Moreover, no causal effect of SA on AD risk was found. Future studies are warranted to investigate cardiovascular pathways between sleep disorders, including SA, and dementia.

## INTRODUCTION

Sleep apnea (SA), a common respiratory disorder in the elderly, has been linked to an increased risk of dementia in numerous epidemiological studies (1–3). However, the types of dementia associated with SA remain uncertain. Some studies have suggested an association between SA and Alzheimer’s disease (AD) (1,4,5), whereas others have highlighted a link with vascular dementia (1,3). Currently, two main mechanistic pathways are hypothesized. Firstly, SA may promote the accumulation of AD proteins such as amyloid-β and tau proteins in the brain (6–8). Secondly, SA may increase the risk of cardiovascular disease (CVD) (9) and CVD risk factors, which are themselves established risk factors for dementia (10,11). However, these hypotheses primarily rely on findings from observational studies which are limited by biases including residual confounding and reverse causality. Moreover, it is difficult to differentiate between neurodegenerative and cerebrovascular pathways since mixed pathology is often more prevalent than pure forms of AD (12), especially with increasing age. Clarifying the causality between SA and AD and CVD might help understanding the biological mechanisms underlying the SA-dementia relationship, which is an important research area given the potential of sleep as a modifiable factor to prevent dementia.

Mendelian randomization (MR) is a method that estimates causal effects by leveraging naturally randomized genetic variation. This approach limits confounding bias due to the random assignment of genes at conception and minimizes reverse causality bias because diseases cannot affect an individual’s germline genetic variation. In the literature, two previous MR studies did not detect a causal effect of SA on AD (13,14), whereas heterogeneous results have been found for SA and CVD outcomes (15–20). These studies were limited by use of older genome-wide association study (GWAS) datasets, low-powered genetic instruments for SA (14), and lack of investigation into potential reverse causal associations (16–20). Furthermore, SA may also be a consequence of AD and CVDs (9,21), and so new approaches are needed to better understand the potential bidirectionality of these relationships. Therefore, our goal was to examine the bidirectional causal relationships between SA and the risks of AD and CVDs (coronary artery disease (CAD) and stroke) by performing MR analyses using the most recent GWAS available.

## METHODS

### Study design and data sources

We conducted this MR study using summary-level data obtained from large, recent, and publicly accessible GWAS (Supplementary Table S1). All GWAS were restricted to European ancestry to minimize potential bias due to population stratification, and ethical approval was granted in original studies.

For the exposure, we obtained GWAS summary statistics (GWAS-SS) from the most recent and largest GWAS on SA (n = 523,366 from five cohorts, including 20,008 SA cases) (22). This GWAS used a multi-trait analysis approach to enhance statistical power, leveraging the high genetic correlations between SA and snoring. SA cases were identified using the International Classification of Diseases (ICD) 9/10 Revision diagnostic codes from electronic health records or self-reported data (either through diagnostic information or answer to the item “stop breathing during sleep”). All cohorts included age, sex, genotype batch (where relevant), and genetic ancestry principal components derived from genotype data as covariates.

Genetic variants association estimates with the risk of late-onset AD, CAD, and stroke were used as the outcomes. For AD, we used GWAS-SS from the largest available GWAS of clinically diagnosed AD, conducted by the International Genomics of Alzheimer’s Project (n = 94,437) (23). For CAD, GWAS-SS were taken from the latest GWAS available combining eight cohorts with the CARDIoGRAMplusC4D consortium (n = 1,165,690) (24). For stroke, we obtained GWAS-SS from the GIGASTROKE consortium, the latest and largest GWAS available (n = 1,308,460) (25) (Supplementary Table S1).

### Selection of instrumental variables

To estimate causal effects, MR analysis uses genetic variants as instrumental variables (IVs), which must satisfy three core assumptions: (i) the IVs should be associated with the exposure (relevance); (ii) the IVs should not be associated with any confounding factors (independence); and (iii) the IVs should affect the outcome solely through their impact on the exposure (exclusion-restriction) (26). Based on these assumptions, we identified IVs as independent single-nucleotide polymorphisms (SNPs) that were significantly associated with SA at a genome-wide level (p-value < 5×10^-8^). To ensure their independence, we excluded duplicate SNPs and performed linkage disequilibrium clumping (r^2^ > 0.001, 10 MB window, using the 1000 Genomes Project as the European reference panel). We calculated the F-statistic for the exposure to evaluate the strength of the IVs, as previously described (27). Then, we extracted these IVs in each of the three outcome GWAS datasets. If a specific SNP was not present, we used a proxy SNP with high linkage disequilibrium (r^2^ > 0.8, using a European reference). To ensure consistency, we harmonized the exposure and outcome GWAS datasets so that the effects corresponded to the same alleles. Finally, we applied additional filtering criteria, removing palindromic and ambiguous SNPs (minor allele frequency >0.42) as well as SNPs with incompatible alleles (26). SNPs showing genome-wide significance for the outcome were also excluded from the analyses (28). For the analyses involving AD, we further excluded variants located ± 250 kb from the *APOE ε4* defining SNP, rs429358, due to its pleiotropic nature which represents a violation of the exclusion-restriction assumption (29).

### Statistical analysis

We conducted two-sample MR analyses to estimate the causal effects of genetically predicted SA on the risk of AD, CAD, and stroke. Fixed-effects inverse variance weighted (IVW) approach was carried out as the primary method. To evaluate if the causal estimates were robust to violations of MR assumptions, diagnostics tests were performed. We employed the MR-Egger regression intercept test to assess for directional horizontal pleiotropy (26), while Cochran’s Q test was used to estimate between-SNP heterogeneity in the estimate of the causal effect. Moreover, the impact of outlier genetic instruments was assessed by two methods: (i) we performed leave-one-out analysis (for IVW and MR-Egger approaches), excluding one IV at a time, to explore the contribution of individual SNPs to the overall effects; and (ii) we conducted radial-MR analysis (“RadialMR” version 1.1 package) to identify data points with large contributions to Cochran’s Q statistic, and we used PhenoScanner (r^2^ > 0.8, using a European reference) to obtain further information on these SNPs. Detected outliers were removed from the analyses. If diagnostics issues were identified, sensitivity analyses using MR-Egger, weighted median, and weighted mode methods were applied. Random-effects IVW method was also performed in a supplementary analysis. Additionally, if significant causal associations were observed, three further sensitivity analyses were carried out. First, to address any potential bias from sample overlap between the exposure and outcome datasets, cross-trait linkage disequilibrium score regression was performed, allowing us to calculate the corrected IVW causal effect estimate using the “MRlap” version 0.0.3 package (30). Second, considering the well-known associations between obesity and SA (31) and the potential strong confounding effect of obesity in the SA-CVD association (Supplementary Figure S1), multivariable MR (MVMR) analysis using the IVW approach was conducted, adjusting for genetically predicted body mass index (BMI) (see Supplementary Table S1 for GWAS-SS details) (32). Third, due to the low statistical power in the MVMR analyses, we also assessed the impact of obesity on the results by excluding the SNPs associated with BMI at a genome-wide level (p-value < 5×10^-8^) in any BMI GWAS dataset. These SNPs were identified via online PhenoScanner. Finally, we explored potential reverse causation by conducting MR analyses in the reverse direction, with the SA phenotype as the outcome and AD and CVDs as the exposures. All statistical analyses were carried out using R version 4.3.0, with the “TwoSampleMR” version 0.5.7 package (26). Codes is publicly accessible online (https://github.com/ccavailles/Sleep-apnea-AD-MR;https://github.com/ccavailles/Sleep-apnea-CAD-MR; https://github.com/ccavailles/Sleep-apnea-stroke-MR).

## RESULTS

We used 32 genetic variants associated with SA as IVs in this MR analysis. In each analysis involving CAD and stroke, one SNP was excluded respectively due to its identification as an outlier. The SNPs used as IVs, their harmonized effects, the identified outliers, and the BMI-associated SNPs are displayed in Supplementary Tables S2 and S3. Supplementary Figures S2 to S7 show the results from leave-one-out and radial-MR analyses. Results using the random-effects IVW approach are presented in Supplementary Table S4 as they were similar to the ones obtained with the fixed-effects IVW method.

### Causal effects between SA and AD

Genetically predicted SA did not influence the risk of AD (odds ratio (OR_IVW_) = 1.14 per log-odds increase in SA liability, 95% confidence interval (CI) = 0.91-1.43; Table 1 and Figure 1). There was no evidence of heterogeneity (Cochran’s Q statistic, p-value = 0.09) or pleiotropy (MR-Egger intercept, p-value = 0.36) effects were observed. In the reverse direction, genetically predicted AD did not influence the risk of SA (OR_IVW_ = 1.01, 95% CI = 0.99-1.02; Table 1 and Figure 2).

**Figure 1.**
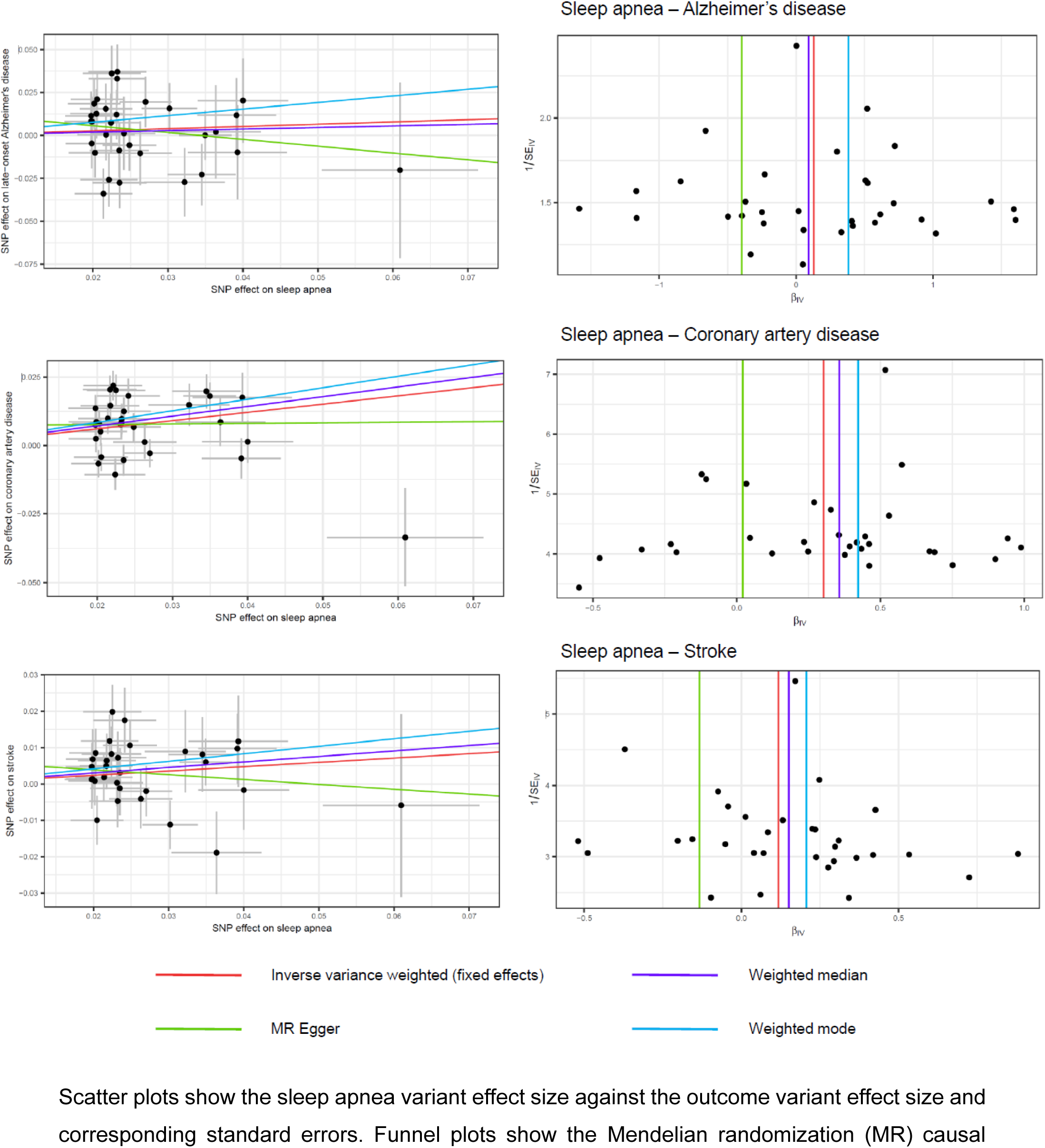
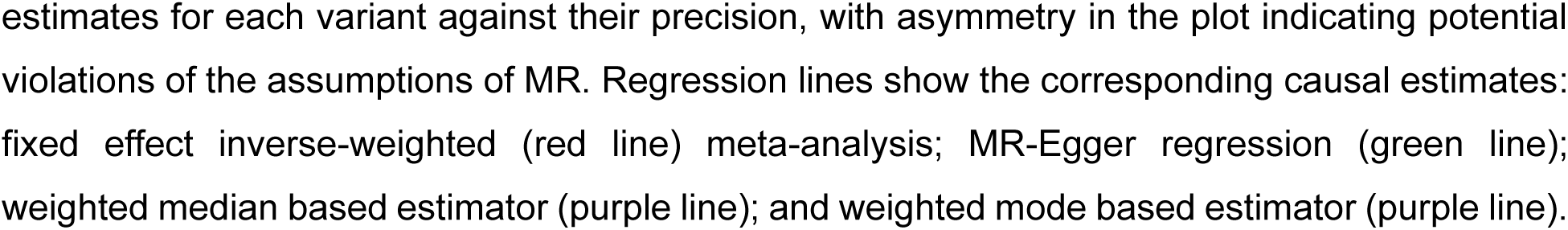
Scatter and funnel plots for each relationship between sleep apnea and the different outcomes (Alzheimer’s disease, coronary artery disease, and stroke).

**Figure 2.**
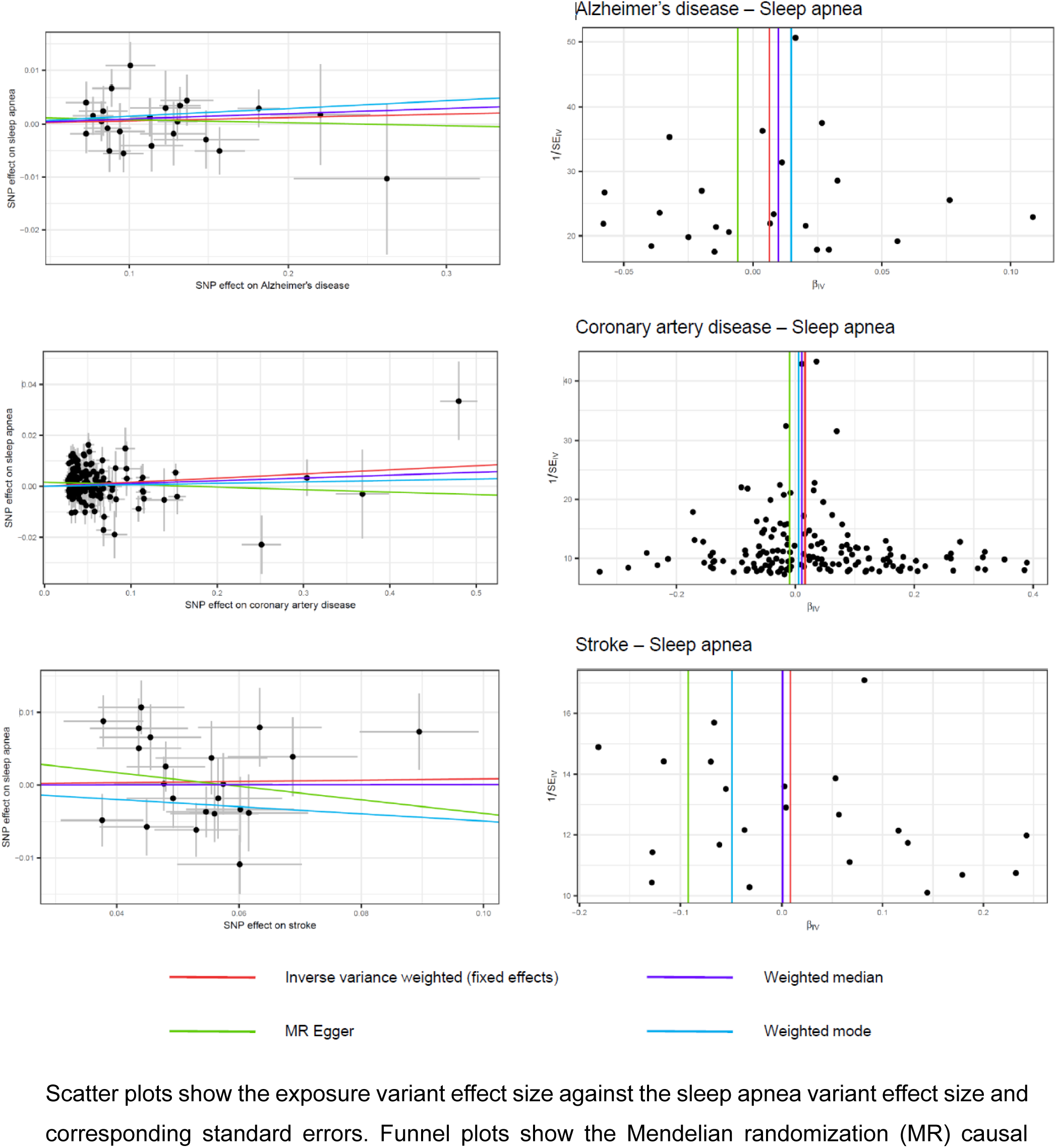
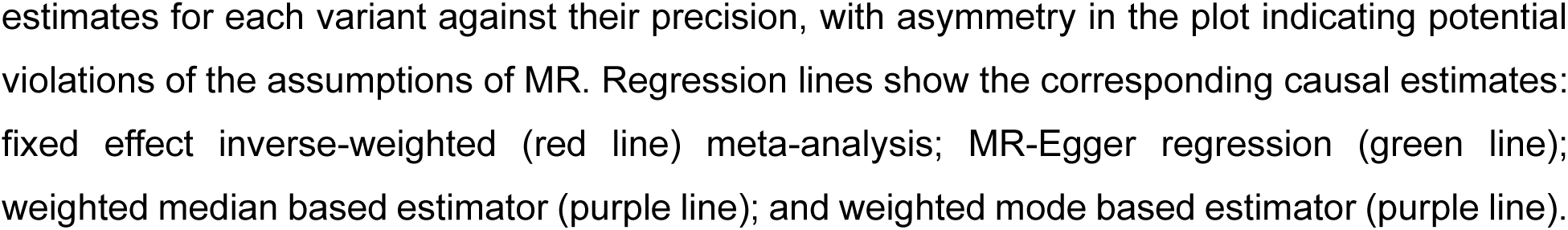
Scatter and funnel plots for each relationship in the bidirectional analysis between the different exposures (Alzheimer’s disease, coronary artery disease, and stroke) and sleep apnea.

**Table 1.** Mendelian randomization estimates for the effect of genetically predicted sleep apnea on the risk of Alzheimer disease, coronary artery disease, and stroke, and their reverse causality.

| Exposure | Outcome | SNP | n | F- | Fixed-effects IVW | MR-Egger | Weighted median | Weighted mode | Cochran's | MR-Egger |
| --- | --- | --- | --- | --- | --- | --- | --- | --- | --- | --- |
|  |  |  | outlier | statistic |  |  |  |  | Q Test | intercept |
|  |  |  |  |  | OR (95%CI) | OR (95%CI) | OR (95%CI) | OR (95%CI) | Q p-value | p-value |
| **Forward** |  |  |  |  |  |  |  |  |  |  |
| SA | AD | 32 | 0 | 41.0 | 1.14 (0.91;1.43) | 0.67 (0.21;2.11) | 1.10 (0.79;1.52) | 1.47 (0.78;2.77) | 0.09 | 0.36 |
| SA | CAD | 31 | 1 | 40.2 | 1.35 (1.25;1.47) | 1.02 (0.58;1.80) | 1.43 (1.25;1.64) | 1.53 (1.16;2.00) | 6.48E-07 | 0.33 |
| SA | Stroke | 31 | 1 | 41.2 | 1.13 (1.01;1.25) | 0.87 (0.55;1.40) | 1.16 (1.01;1.34) | 1.23 (0.93;1.62) | 0.43 | 0.29 |
| **Reverse** |  |  |  |  |  |  |  |  |  |  |
| AD | SA | 23 | 0 | 56.1 | 1.01 (0.99;1.02) | 0.99 (0.94;1.05) | 1.01 (0.99;1.03) | 1.01 (0.98;1.05) | 0.51 | 0.64 |
| CAD | SA | 159 | 4 | 79.2 | 1.02 (1.00;1.03) | 0.99 (0.96;1.02) | 1.01 (0.99;1.03) | 1.01 (0.98;1.03) | 1.62E-07 | 0.06 |
| Stroke | SA | 22 | 1 | 44.0 | 1.01 (0.98;1.04) | 0.91 (0.71;1.17) | 1.00 (0.95;1.05) | 0.95 (0.85;1.06) | 0.001 | 0.43 |

Abbreviations: AD, Alzheimer's disease; CAD, coronary artery disease; CI, confidence interval; IVW, inverse variance weighted; OR, odds ratio; SA, sleep apnea; SNP, single nucleotide polymorphism

### Causal effects between SA and CAD

Genetically predicted SA was associated with higher risk of CAD (OR_IVW_ = 1.35, 95% CI = 1.25-1.47; Table 1 and Figure 1). Heterogeneity was detected (Figure 1), but MR sensitivity analyses were significant, except for the MR-Egger estimate, and were consistent in effect direction. A significant difference between observed and corrected effects was found in the analysis correcting for sample overlap. After correction, genetically predicted SA was still significantly associated with higher risk of CAD, although the estimate was somewhat attenuated (OR_IVW-corrected_ = 1.13, 95% CI = 1.06-1.20). In the MVMR analysis adjusting for BMI, the causal relationship became nonsignificant (OR_IVW_ = 1.05, 95% CI = 0.92-1.21; Supplementary Table S5). However, this specific analysis was limited by weak instruments (conditional F-statistics <10) which represents a violation of the relevance MR assumption. Therefore, we explored the impact of excluding the BMI-associated SNPs on the results and found that genetically predicted SA still significantly increased the risk of CAD (OR_IVW_ = 1.26, 95% CI = 1.15-1.39; Supplementary Table S6). In the bidirectional analysis, a significant causal effect of genetically predicted CAD on SA risk was found using the IVW approach (OR_IVW_ = 1.02, 95% CI = 1.00-1.03; Table 1 and Figure 2). However, there was evidence of heterogeneity, and all the sensitivity analyses were nonsignificant, suggesting a potential bias in the IVW causal estimate.

### Causal effects between SA and stroke

We found a significant causal effect of genetically predicted SA on stroke (OR_IVW_ = 1.13, 95% CI = 1.01-1.25; Table 1 and Figure 1); there was no evidence of heterogeneity (Cochran’s Q statistic, p-value = 0.43) or pleiotropy as evidenced by the Egger intercept (p-value = 0.29). Analysis correcting for sample overlap did not reveal a significant difference between observed and corrected effects, suggesting that the IVW estimate are not biased by sample overlap. In the MVMR analysis adjusting for BMI, the causal relationship became non-significant (OR_IVW_ = 1.03, 95% CI = 0.91-1.16; Supplementary Table S5) suggesting that BMI could confound the association; but this analysis was limited by weak instruments (conditional F-statistics <10). After excluding the BMI-associated SNPs, the causal effect also became non-significant (OR_IVW_ = 1.08, 95% CI = 0.96-1.22; Supplementary Table S6). Finally, the bidirectional MR analysis indicated no causal effect for genetically predicted stroke on the risk of SA (OR_IVW_ = 1.01, 95% CI = 0.98-1.04; Table 1 and Figure 2).

## DISCUSSION

Using the most recent GWAS datasets available, this MR study revealed that genetically predicted SA increased the risk of CAD, while the causal association with stroke risk may be confounded by BMI. Furthermore, findings do not support evidence of a causal link between genetically predicted SA and AD risk. In the bidirectional analyses, no causal effects were observed for genetically predicted AD, CAD, or stroke on the risk of SA. Taken together, these findings suggest that cerebrovascular pathology may play a more important role than AD pathology in the relationship between SA and dementia.

Numerous observational studies have established a link between SA and an increased risk of cognitive impairment and all-cause dementia (1–3,33). However, it remains controversial which type of dementia is driving this association. Some studies have found an association between SA and AD (1,4,5), whereas others have not (34). Moreover, very few studies have investigated the association between SA and vascular dementia, also reporting conflicting findings (1,3,35). These discrepancies might be due in part to the limitations of observational studies which are more prone to several sources of bias (e.g., confounders bias and reverse causality). In this study, we used a MR approach to overcome these limitations. We did not yield evidence supporting a causal effect of SA on AD, aligning with the results of the two previous MR studies examining this causal relationship (13,14). However, our findings suggest that cerebrovascular pathology would be a more important pathway in the SA-dementia relationship. This is consistent with the well-established vascular risk factors of dementia (10,11,36) as well as the vascular consequences of SA (9).

Notably, most observational studies, but not all (37,38), have reported an association between SA and increased risk of CAD and stroke (39–41). In contrast, previous MR studies did not establish a causal effect of SA on stroke risk (15–18), whereas results for CAD were mixed (15,16,19,20). Specifically, SA did not increase the risk of CAD in two MR studies (16,19), while a suggestive association was observed in two other studies (15,20). Our findings contribute to the literature by highlighting a strong causal relationship between genetically predicted SA and higher risk of CAD, and by showing that the causal association with stroke risk was confounded by BMI. These differences may be attributable to the use of smaller GWAS datasets for the exposure and/or outcomes in previous MR studies, along with a limited number of valid IVs.

Given the strong genetic correlation between SA and obesity, accounting for BMI is important as their pathways leading to CVDs may be confounded. Indeed, the role of BMI in the SA-CVDs associations remains controversial. In observational studies, some research has shown associations between SA, CAD, and stroke independently of BMI (37,38,40), while some others have not (37,41,42). Similarly, we found that genetically predicted SA increased the risk of CAD independently of BMI, while the causal effect of genetically predicted SA on stroke risk was confounded by BMI. Further studies with higher statistical power are warranted to replicate these results. Although we do not have a clear explanation for these differences, our results primarily hallmark the important role of BMI and suggest that it may explain the entirety (e.g., stroke risk) or only a part (e.g., CAD) of the SA-CVD association. SA might impact CVDs through several mechanisms including, but not limited to, intermittent hypoxia, oxidative stress, inflammation, endothelial dysfunction, white matter lesions, and atherosclerosis (2,9).

Overall, these findings suggest a greater role for cerebrovascular pathology than AD pathology in the relationship between SA and dementia. This observation aligns with mounting evidence involving vascular damage, such as infarcts and white matter changes, as a common feature in various types of dementia (12,43). It il also consistent with the importance of vascular cognitive impairment and dementia (44), underscoring the complex interplay between neurodegenerative and vascular mechanisms. Future studies should investigate these causal relationships using amyloid/tau and cerebrovascular phenotypes rather than clinical phenotypes.

While it remains possible that both pathologies contribute to dementia in varying degrees, addressing vascular risk factors and SA through lifestyle modifications and medical interventions may be an important strategy in reducing the risk of dementia (45,46).

Strengths of our study include a bidirectional MR approach allowing a better understanding of the direction of the causal effects, the use of large-scale GWAS-SS, a small magnitude of weak instrument bias in the main analyses (F-statistics of the IVs were greater than 10 for all exposures), and multiple sensitivity analyses to confirm the robustness of the results. However, our findings should be interpreted in light of several limitations. First, considering SA is a binary exposure, our estimates represent the average causal effects in “compliers” (i.e., individuals for whom SA would be present if they have the genetic variant and absent otherwise) (47). Therefore, estimates should be interpreted as the effect of liability to SA on the outcome, rather than exact causal effect. Second, since SA was evaluated from primary care records or self-reported data, underdiagnosis is possible which might bias the results of the associations between the genetic variants and SA towards the nulls. Third, potential bias toward observational associations might be present when the exposure and the outcome datasets overlap (48). To address this, we performed a cross-trait linkage disequilibrium score regression analysis to verify the reliability of the identified causal effect of SA on CAD and stroke risks (around 20% overlap for both datasets), and results remained unchanged. Fourth, despite the use of the largest and more recent GWAS datasets available, we didn’t have enough statistical power to report robust conclusions in the MVMR analyses adjusting for BMI. Further studies are needed to decipher the potential mediating role of BMI in the SA-CVDs associations. In addition, we were not able to directly assess vascular dementia as no sufficiently robust GWAS has been published to date. Fifth, competing risks with death and other CVDs cannot be excluded and may lead to false null findings. This limitation is particularly relevant for late onset diseases such as AD. Future studies are thus warranted to confirm the current results. Finally, we restricted our analyses to European-ancestry participants which might limit the generalizability of our findings to other populations.

Among individuals of European ancestry, this MR study supports the hypothesis that genetically predicted SA increased the risk of CAD, whereas the causal effect on stroke risk was confounded by BMI. Furthermore, genetically predicted SA may not have a causal effect on the development of AD. These findings may prompt subsequent investigations aimed at exploring therapeutic approaches targeting SA to prevent CVD risks (49,50), while also elucidating the role of BMI in these associations. Furthermore, they could lead to additional research investigating cardiovascular mediating pathways between sleep and dementia development, thereby enhancing our comprehension of the biological mechanisms that underlie this association.

## ACKNOWLEDGMENTS

The authors thank the investigators from all the GWAS used in this study as well as the research participants.

## DATA AVAILABILITY

This study used summary results from published research papers, with the references for those studies provided in the main manuscript. The sleep apnea data are available on request after approval by Dr Campos and Dr Renteria. Coronary artery disease, stroke, and body mass index data are publicly available. Supplementary Tables S2 and S3 provide the harmonized SNP effects needed to reproduce the results of this analysis. The codes used to conduct this analysis are publicly available online at: https://github.com/ccavailles/Sleep-apnea-AD-MR; https://github.com/ccavailles/Sleep-apnea-stroke-MR; https://github.com/ccavailles/Sleep-apnea-CAD-MR).

## SUPPLEMENTAL MATERIAL

Tables S1-S6

Figures S1-S7

## REFERENCES

1. Yaffe K, Nettiksimmons J, Yesavage J, Byers A. Sleep Quality and Risk of Dementia Among Older Male Veterans. Am J Geriatr Psychiatry. 2015 Feb 1;23.

2. Yaffe K, Laffan AM, Harrison SL, Redline S, Spira AP, Ensrud KE, Ancoli-Israel S, Stone KL. Sleep-disordered breathing, hypoxia, and risk of mild cognitive impairment and dementia in older women. JAMA. 2011 Aug 10;306(6):613–9.

3. Chang WP, Liu ME, Chang WC, Yang AC, Ku YC, Pai JT, Huang HL, Tsai SJ. Sleep Apnea and the Risk of Dementia: A Population-Based 5-Year Follow-Up Study in Taiwan. PLOS ONE. 2013 Oct 24;8(10):e78655.

4. Lee JE, Yang SW, Ju YJ, Ki SK, Chun KH. Sleep-disordered breathing and Alzheimer’s disease: A nationwide cohort study. Psychiatry Res. 2019 Mar 1;273:624–30.

5. Tsai MS, Li HY, Huang CG, Wang RYL, Chuang LP, Chen NH, Liu CH, Yang YH, Liu CY, Hsu CM, et al. Risk of Alzheimer’s Disease in Obstructive Sleep Apnea Patients With or Without Treatment: Real-World Evidence. The Laryngoscope. 2020 Sep;130(9):2292–8.

6. André C, Rehel S, Kuhn E, Landeau B, Moulinet I, Touron E, Ourry V, Le Du G, Mézenge F, Tomadesso C, et al. Association of Sleep-Disordered Breathing With Alzheimer Disease Biomarkers in Community-Dwelling Older Adults: A Secondary Analysis of a Randomized Clinical Trial. JAMA Neurol. 2020 Jun 1;77(6):716–24.

7. Sharma RA, Varga AW, Bubu OM, Pirraglia E, Kam K, Parekh A, Wohlleber M, Miller MD, Andrade A, Lewis C, et al. Obstructive Sleep Apnea Severity Affects Amyloid Burden in Cognitively Normal Elderly. A Longitudinal Study. Am J Respir Crit Care Med. 2018 Apr 1;197(7):933–43.

8. Yaffe K, Falvey CM, Hoang T. Connections between sleep and cognition in older adults. Lancet Neurol. 2014 Oct;13(10):1017–28.

9. Yeghiazarians Y, Jneid H, Tietjens JR, Redline S, Brown DL, El-Sherif N, Mehra R, Bozkurt B, Ndumele CE, Somers VK. Obstructive Sleep Apnea and Cardiovascular Disease: A Scientific Statement From the American Heart Association. Circulation. 2021 Jul 20;144(3):e56–67.

10. Kuźma E, Lourida I, Moore SF, Levine DA, Ukoumunne OC, Llewellyn DJ. Stroke and dementia risk: A systematic review and meta-analysis. Alzheimers Dement. 2018 Nov;14(11):1416–26.

11. Deckers K, Schievink SHJ, Rodriquez MMF, van Oostenbrugge RJ, van Boxtel MPJ, Verhey FRJ, Köhler S. Coronary heart disease and risk for cognitive impairment or dementia: Systematic review and meta-analysis. PLoS ONE. 2017 Sep 8;12(9):e0184244.

12. Kapasi A, DeCarli C, Schneider JA. Impact of multiple pathologies on the threshold for clinically overt dementia. Acta Neuropathol. 2017 Aug;134(2):171–86.

13. Li J, Zhao L, Ding X, Cui X, Qi L, Chen Y. Obstructive sleep apnea and the risk of Alzheimer’s disease and Parkinson disease: A Mendelian randomization study OSA, Alzheimer’s disease and Parkinson disease. Sleep Med. 2022 Sep;97:55–63.

14. Cullell N, Cárcel-Márquez J, Gallego-Fábrega C, Muiño E, Llucià-Carol L, Lledós M, Amaut KEU, Krupinski J, Fernández-Cadenas I. Sleep/wake cycle alterations as a cause of neurodegenerative diseases: A Mendelian randomization study. Neurobiol Aging. 2021 Oct;106:320.e1–320.e12.

15. Wang J, Campos AI, Rentería ME, Xu L. Causal associations of sleep apnea, snoring with cardiovascular diseases, and the role of body mass index: a two-sample Mendelian randomization study. Eur J Prev Cardiol. 2023 May 9;30(7):552–60.

16. Li Y, Miao Y, Zhang Q. Causal associations of obstructive sleep apnea with cardiovascular disease: a Mendelian randomization study. Sleep. 2023 Mar 1;46(3):zsac298.

17. Li P, Dong Z, Chen W, Yang G. Causal Relations Between Obstructive Sleep Apnea and Stroke: A Mendelian Randomization Study. Nat Sci Sleep. 2023 Apr 19;15:257–66.

18. Titova OE, Yuan S, Baron JA, Lindberg E, Michaëlsson K, Larsson SC. Sleep-disordered breathing-related symptoms and risk of stroke: cohort study and Mendelian randomization analysis. J Neurol. 2022 May 1;269(5):2460–8.

19. Ding X, Zhao L, Cui X, Qi L, Chen Y. Mendelian randomization reveals no associations of genetically-predicted obstructive sleep apnea with the risk of type 2 diabetes, nonalcoholic fatty liver disease, and coronary heart disease. Front Psychiatry. 2023 Feb 9;14:1068756.

20. Titova OE, Yuan S, Baron JA, Lindberg E, Michaëlsson K, Larsson SC. Self-reported symptoms of sleep-disordered breathing and risk of cardiovascular diseases: Observational and Mendelian randomization findings. J Sleep Res. 2022;31(6):e13681.

21. Emamian F, Khazaie H, Tahmasian M, Leschziner GD, Morrell MJ, Hsiung GYR, Rosenzweig I, Sepehry AA. The Association Between Obstructive Sleep Apnea and Alzheimer’s Disease: A Meta-Analysis Perspective. Front Aging Neurosci. 2016 Apr 12;8:78.

22. Campos AI, Ingold N, Huang Y, Mitchell BL, Kho PF, Han X, García-Marín LM, Ong JS, 23andMe Research Team, Law MH, et al. Discovery of genomic loci associated with sleep apnea risk through multi-trait GWAS analysis with snoring. Sleep. 2023 Mar 9;46(3):zsac308.

23. Kunkle BW, Grenier-Boley B, Sims R, Bis JC, Damotte V, Naj AC, Boland A, Vronskaya M, Van der Lee SJ, Amlie-Wolf A, et al. Genetic meta-analysis of diagnosed Alzheimer’s disease identifies new risk loci and implicates Aβ, tau, immunity and lipid processing. Nat Genet. 2019 Mar;51(3):414–30.

24. Aragam KG, Jiang T, Goel A, Kanoni S, Wolford BN, Atri DS, Weeks EM, Wang M, Hindy G, Zhou W, et al. Discovery and systematic characterization of risk variants and genes for coronary artery disease in over a million participants. Nat Genet. 2022 Dec;54(12):1803–15.

25. Mishra A, Malik R, Hachiya T, Jürgenson T, Namba S, Posner DC, Kamanu FK, Koido M, Le Grand Q, Shi M, et al. Stroke genetics informs drug discovery and risk prediction across ancestries. Nature. 2022 Nov;611(7934):115–23.

26. Hemani G, Zheng J, Elsworth B, Wade KH, Haberland V, Baird D, Laurin C, Burgess S, Bowden J, Langdon R, et al. The MR-Base platform supports systematic causal inference across the human phenome. Elife. 2018 May 30;7:e34408.

27. Burgess S, Thompson SG, CRP CHD Genetics Collaboration. Avoiding bias from weak instruments in Mendelian randomization studies. Int J Epidemiol. 2011 Jun 1;40(3):755–64.

28. Thompson SB Simon G. Mendelian Randomization: Methods for Causal Inference Using Genetic Variants. 2nd ed. New York: Chapman and Hall/CRC; 2021. 240 p.

29. Watanabe K, Stringer S, Frei O, Umićević Mirkov M, de Leeuw C, Polderman TJC, van der Sluis S, Andreassen OA, Neale BM, Posthuma D. A global overview of pleiotropy and genetic architecture in complex traits. Nat Genet. 2019 Sep;51(9):1339–48.

30. Mounier N, Kutalik Z. Bias correction for inverse variance weighting Mendelian randomization. Genet Epidemiol. 2023;47(4):314–31.

31. Kuvat N, Tanriverdi H, Armutcu F. The relationship between obstructive sleep apnea syndrome and obesity: A new perspective on the pathogenesis in terms of organ crosstalk. Clin Respir J. 2020;14(7):595–604.

32. Yengo L, Sidorenko J, Kemper KE, Zheng Z, Wood AR, Weedon MN, Frayling TM, Hirschhorn J, Yang J, Visscher PM, et al. Meta-analysis of genome-wide association studies for height and body mass index in ∼700000 individuals of European ancestry. Hum Mol Genet. 2018 Oct 15;27(20):3641–9.

33. Leng Y, McEvoy CT, Allen IE, Yaffe K. Association of Sleep-Disordered Breathing With Cognitive Function and Risk of Cognitive Impairment: A Systematic Review and Meta-analysis. JAMA Neurol. 2017 Oct 1;74(10):1237–45.

34. Lutsey PL, Misialek JR, Mosley TH, Gottesman RF, Punjabi NM, Shahar E, MacLehose R, Ogilvie RP, Knopman D, Alonso A. Sleep characteristics and risk of dementia and Alzheimer’s disease: The Atherosclerosis Risk in Communities Study. Alzheimers Dement. 2018 Feb;14(2):157–66.

35. Elwood P, Bayer A, Fish M, Pickering J, Mitchell C, Gallacher J. Sleep disturbance and daytime sleepiness predict vascular dementia. J Epidemiol Community Health. 2011 Sep 1;65:820–4.

36. Livingston G, Huntley J, Sommerlad A, Ames D, Ballard C, Banerjee S, Brayne C, Burns A, Cohen-Mansfield J, Copper C, et al. Dementia prevention, intervention, and care: 2020 report of the Lancet Commission. The Lancet. 2020 Aug 8;396(10248):413–46.

37. Catalan-Serra P, Campos-Rodriguez F, Reyes-Nuñez N, Selma-Ferrer MJ, Navarro-Soriano C, Ballester-Canelles M, Soler-Cataluña JJ, Roman-Sanchez P, Almeida-Gonzalez CV, Martinez-Garcia MA. Increased Incidence of Stroke, but Not Coronary Heart Disease, in Elderly Patients With Sleep Apnea. Stroke. 2019 Feb;50(2):491–4.

38. Marshall NS, Wong KKH, Cullen SRJ, Knuiman MW, Grunstein RR. Sleep Apnea and 20- Year Follow-Up for All-Cause Mortality, Stroke, and Cancer Incidence and Mortality in the Busselton Health Study Cohort. J Clin Sleep Med. 2014 Apr 15;10(4):355–62.

39. Carthy CEM, Yusuf S, Judge C, Alvarez-Iglesias A, Hankey GJ, Oveisgharan S, Damasceno A, Iversen HK, Rosengren A, Avezum A, et al. Sleep Patterns and the Risk of Acute Stroke: Results From the INTERSTROKE International Case-Control Study. Neurology. 2023 May 23;100(21):e2191–203.

40. Strausz S, Havulinna AS, Tuomi T, Bachour A, Groop L, Mäkitie A, Koskinen S, Salomaa V, Palotie A, Ripatti S, et al. Obstructive sleep apnoea and the risk for coronary heart disease and type 2 diabetes: a longitudinal population-based study in Finland. BMJ Open. 2018 Oct 15;8(10):e022752.

41. Redline S, Yenokyan G, Gottlieb DJ, Shahar E, O’Connor GT, Resnick HE, Diener-West M, Sanders MH, Wolf PA, Geraghty EM, et al. Obstructive Sleep Apnea–Hypopnea and Incident Stroke. Am J Respir Crit Care Med. 2010 Jul 15;182(2):269–77.

42. Gottlieb DJ, Yenokyan G, Newman AB, O’Connor GT, Punjabi NM, Quan SF, Redline S, Resnick HE, Tong EK, Diener-West M, et al. A Prospective Study of Obstructive Sleep Apnea and Incident Coronary Heart Disease and Heart Failure: The Sleep Heart Health Study. Circulation. 2010 Jul 27;122(4):352–60.

43. Querfurth HW, LaFerla FM. Alzheimer’s disease. N Engl J Med. 2010 Jan 28;362(4):329–44.

44. Wong EC, Chui HC. Vascular Cognitive Impairment and Dementia. Contin Minneap Minn. 2022 Jun 1;28(3):750–80.

45. Dunietz GL, Chervin RD, Burke JF, Conceicao AS, Braley TJ. Obstructive sleep apnea treatment and dementia risk in older adults. Sleep. 2021 Sep 13;44(9):zsab076.

46. Rosenberg A, Ngandu T, Rusanen M, Antikainen R, Bäckman L, Havulinna S, Hänninen T, Laatikainen T, Lehtisalo J, Levälahti E, et al. Multidomain lifestyle intervention benefits a large elderly population at risk for cognitive decline and dementia regardless of baseline characteristics: The FINGER trial. Alzheimers Dement. 2018;14(3):263–70.

47. Burgess S, Labrecque JA. Mendelian randomization with a binary exposure variable: interpretation and presentation of causal estimates. Eur J Epidemiol. 2018;33(10):947–52.

48. Burgess S, Davies NM, Thompson SG. Bias due to participant overlap in two-sample Mendelian randomization. Genet Epidemiol. 2016 Nov;40(7):597–608.

49. Brown DL, Durkalski V, Durmer JS, Broderick JP, Zahuranec DB, Levine DA, Anderson CS, Bravata DM, Yaggi HK, Morgensstern LB, et al. Sleep for Stroke Management and Recovery Trial (Sleep SMART): Rationale and methods. Int J Stroke. 2020 Oct;15(8):923–9.

50. McEvoy RD, Antic NA, Heeley E, Luo Y, Ou Q, Zhang X, Mediano O, Chen R, Drager LF, Liu Z, et al. CPAP for Prevention of Cardiovascular Events in Obstructive Sleep Apnea. N Engl J Med. 2016 Sep 8;375(10):919–31.

