## Supplementary Materials for "Causal Associations of Sleep Apnea with Alzheimer’s Disease and Cardiovascular Disease: a Bidirectional Mendelian Randomization Analysis"

### Supplementary material

**Supplementary Table S1. Description of GWAS datasets used in this study.**

| Study | Trait | Definition | Cohort/consortium | n | Mean age | Females, % |
|---|---|---|---|---|---|---|
| Campos et al, 2023 (22) | Sleep apnea | ICD diagnostic from electronic health records (ICD-9: 327.23 and ICD-10: G47.3), self-reported diagnostic or answer to the item "stop breathing during sleep" | UK Biobank; CLSA; AGDS; Partner's Healthcare Biobank; FinnGen | 351,316 | - | - |
| Kunkle et al, 2019 (23) | Late-onset Alzheimer disease | Clinical diagnostic according to NINCDS-ADRDA/DSM-IV or V or III-R criteria by neurologists or a clinical consensus using various types of data (Clinical Dementia Rating scale, Hachinski Ischemic Score, clinical data, cognitive tests, MRI data, etc.) | IGAP | 94,437 | About 72 years old | About 59% |
| Aragam et al, 2022 (24) | Coronary artery disease | Prevalent or incident CAD, acute coronary syndrome or left main CAD, myocardial infarction, chronic IHD, IHD death, and/or angina; made according to standard criteria (e.g., ICD-9 or 10) using general practice, nursing home records, death certificates, and/or self-reported data | CARDIoGRAMplusC4D; UK Biobank; deCODE Genetics; EPIC-CVD; TIMI clinical trials; GerMIFS; HUNT; GCC; Mass General Brigham Biobank | 1,165,690 | About 53 years old | About 51% |
| Mishra et al, 2022 (25) | Stroke | Any stroke, ischemic stroke, cardioembolic stroke, small vessel stroke, large artery stroke, and/or intracerebral hemorrhage; made according to standard criteria (e.g., ICD-9 or 10, TOAST, WHO) using general practice, nursing home records, death certificates, and/or self-reported data | GIGASTROKE | 1,308,460 | About 57 years old | About 54% |
| Yengo et al, 2018 (32) | Body mass index | Measured or self-reported weight in kg per height in meters squared | UK Biobank, GIANT | 690,495 | - | - |

AGDS, Australian Genetics of Depression Study; CLSA, Canadian Longitudinal Study of Aging; DSM, Diagnostic and Statistical Manual of Mental Disorders; EPIC-CVD, European Prospective Investigation into Cancer and Nutrition – cardiovascular disease; FinnGen, Finland genetic research; GCC, Greek Coronary Disease Cohort; GerMIFS, German Myocardial Infarction Family Study; GIANT, Genetic Investigation of Anthropometric Traits; HUNT, Nord-Trøndelag Health Study; IHD, Ischemic Heart Disease, IGAP, International Genomics of Alzheimer's Project; NINCDS-ADRDA, Neurological and Communicative Diseases and Stroke/Alzheimer's Disease and Related Disorders Association; TIMI, Thrombolysis in Myocardial Infarction; TOAST, Trial of Org 10172 in Acute Stroke Treatment.

**Supplementary Table S4. Mendelian randomization estimates using the random-effects approach for the effect of genetically predicted sleep apnea on the risk of Alzheimer disease, coronary artery disease, and stroke, and their reverse causality.**

| Exposure | Outcome | SNP | n outlier | F-statisitcs | Random- effects inverse-variance weighted OR (95%CI) |
|---|---|---|---|---|---|
| **Forward** | | | | | |
| SA | AD | 32 | 0 | 41.0 | 1.14 (0.88;1.48) |
| SA | CAD | 31 | 1 | 40.2 | 1.35 (1.19;1.54) |
| SA | Stroke | 31 | 1 | 41.2 | 1.13 (1.01;1.25) |
| **Reverse** | | | | | |
| AD | SA | 23 | 0 | 56.1 | 1.01 (0.99;1.02) |
| CAD | SA | 159 | 4 | 79.2 | 1.02 (1.00;1.03) |
| Stroke | SA | 22 | 1 | 44.0 | 1.01 (0.96;1.06) |

Abbreviations: AD, Alzheimer's disease; CAD, coronary artery disease; CI, confidence interval; OR, odds ratio; SNP, single nucleotide polymorphism

**Supplementary Table S5. Multivariable Mendelian randomization associations of coronary artery disease and stroke with sleep apnea and body mass index.**

| | | SNP | F-statisitcs | Fixed-effects inverse-variance weighted | MR-Egger | Weighted median |
|---|---|---|---|---|---|---|
| Exposure | Outcome | | | OR (95%CI) | OR (95%CI) | OR (95%CI) |
| SA | CAD | 10 | 2.4 | 1.05 (0.92;1.21) | 1.05 (0.88;1.25) | 1.02 (0.91;1.16) |
| SA | Stroke | 11 | 2.6 | 1.03 (0.92;1.16) | 1.07 (0.92;1.25) | 1.07 (0.94;1.23) |
| BMI | CAD | 490 | 4.8 | 1.46 (1.35;1.57) | 1.46 (1.34;1.58) | 1.51 (1.40;1.62) |
| BMI | Stroke | 491 | 5.2 | 1.15 (1.08;1.22) | 1.16 (1.07;1.26) | 1.15 (1.06;1.24) |

Abbreviations: AD, Alzheimer’s disease; BMI, body mass index; CAD, coronary artery disease; CI, confidence interval; OR, odds ratio; SNP, single nucleotide polymorphism

Note: We decided not to include the Weighted Mode due to the novelty of the MVMRMode package, which necessitates additional testing and validation.

**Supplementary Table S6. Mendelian randomization estimates for the effect of genetically predicted sleep apnea on the risk of coronary artery disease and stroke after excluding the SNPs associated with body mass index.**

| | | SNP | n outlier | F-statisitcs | Fixed-effects inverse-variance weighted | MR-Egger | Weighted median | Weighted mode | Cochran's Q Test | MR-Egger intercept |
|---|---|---|---|---|---|---|---|---|---|---|
| Exposure | Outcome | | | | OR (95%CI) | OR (95%CI) | OR (95%CI) | OR (95%CI) | Q p-value | p-value |
| SA | CAD | 23 | 2 | 37.5 | 1.26 (1.15;1.39) | 0.72 (0.38;1.36) | 1.28 (1.09;1.50) | 1.44 (1.01;2.06) | 1.4E-5 | 0.09 |
| SA | Stroke | 25 | 1 | 38.6 | 1.08 (0.96;1.22) | 0.75 (0.44;1.29) | 1.09 (0.93;1.29) | 1.26 (0.87;1.83) | 0.36 | 0.18 |

Abbreviations: CAD, coronary artery disease; CI, confidence interval; OR, odds ratio; SA, sleep apnea; SNP, single nucleotide polymorphism

**Supplementary Figure S1. Directed acyclic graph representing the relationships between sleep apnea, cardiovascular disease, and body mass index.**

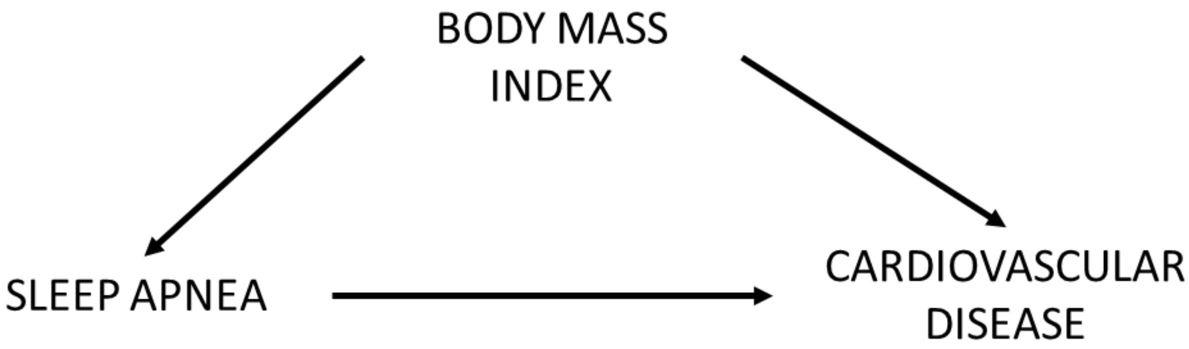

**Supplementary Figure S2. Leave-one-out analysis and radial plots for the relationship between sleep apnea (SA) and Alzheimer's disease (AD) using the fixed-effects inverse-variance weighted approach.**

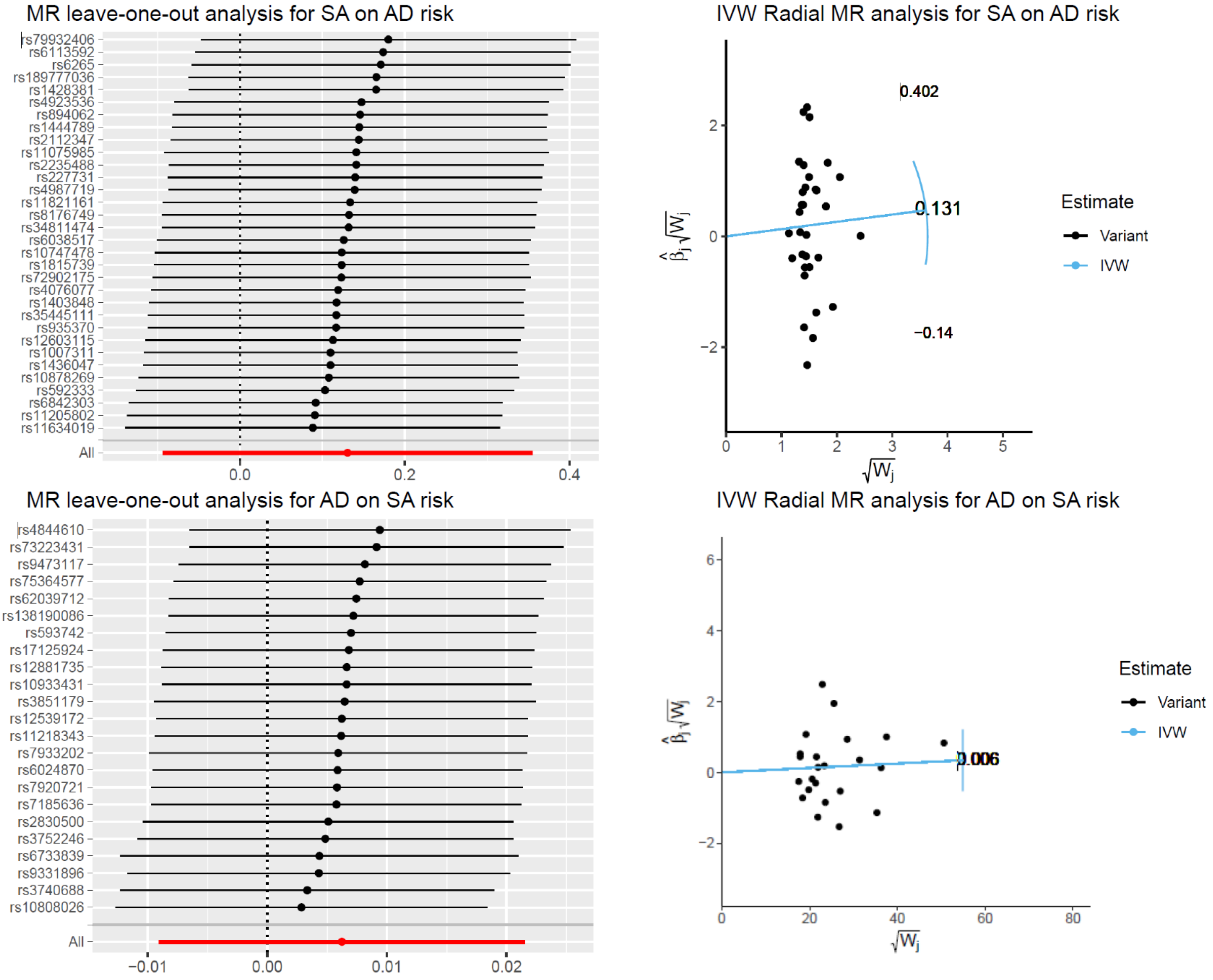


Leave-one-out analysis plots show the Mendelian randomization causal estimates by sequentially dropping one variant at a time, with asymmetry in the plot indicating potential outliers. In the radial plots, the regression line (blue line) shows the radial causal estimate for inverse weighted regression models.

**Supplementary Figure S3. Leave-one-out analysis and radial plots for the relationship between sleep apnea (SA) and stroke using the fixed-effects inverse-variance weighted approach.**

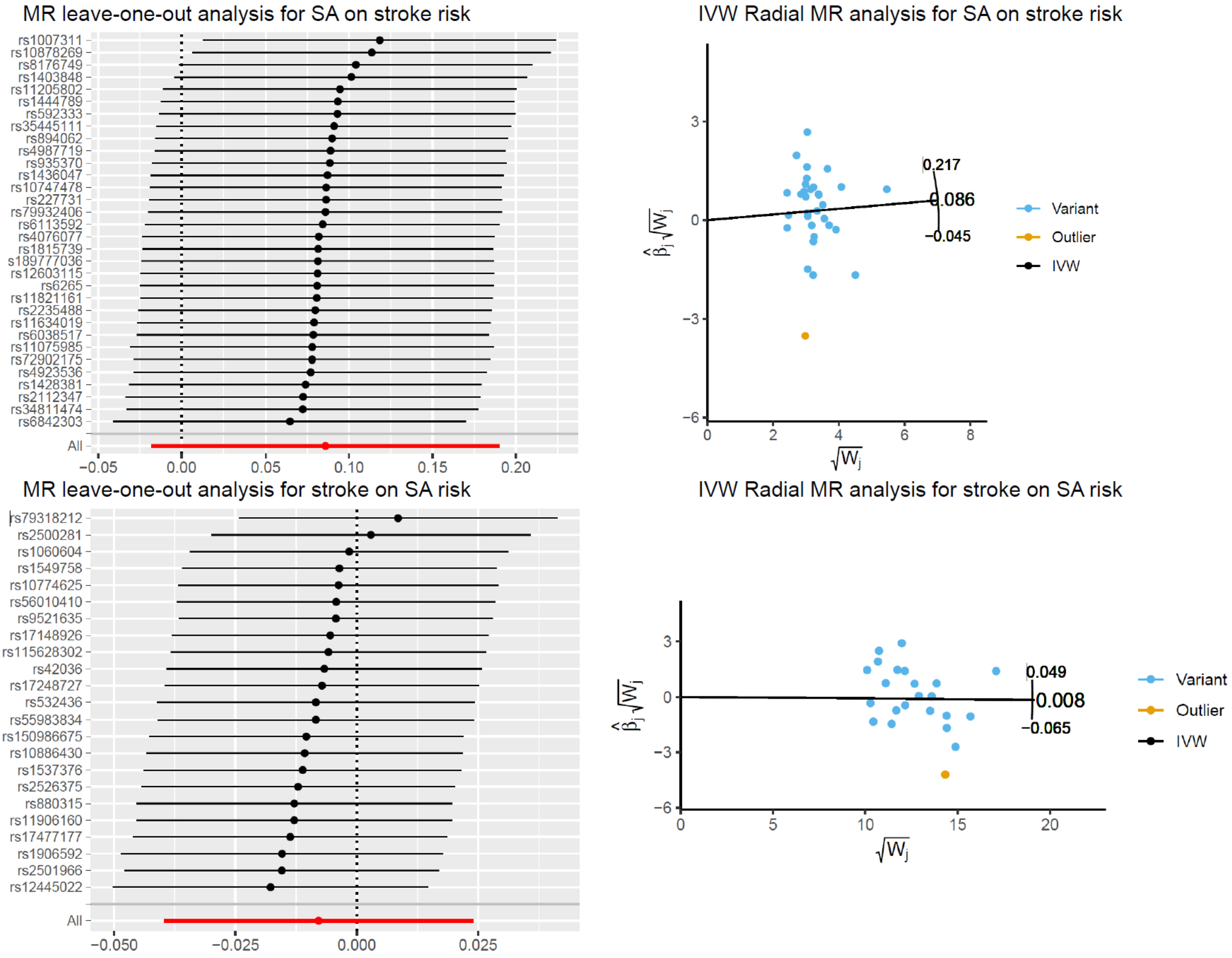


Leave-one-out analysis plots show the Mendelian randomization causal estimates by sequentially dropping one variant at a time, with asymmetry in the plot indicating potential outliers. In the radial plots, the regression line (black line) shows the radial causal estimate for inverse weighted regression models. Variants highlighted in orange were flagged as outliers.

**Supplementary Figure S4. Leave-one-out analysis and radial plot for the effect of sleep apnea (SA) on the risk of coronary artery disease (CAD) using the fixed-effects inverse-variance weighted approach.**

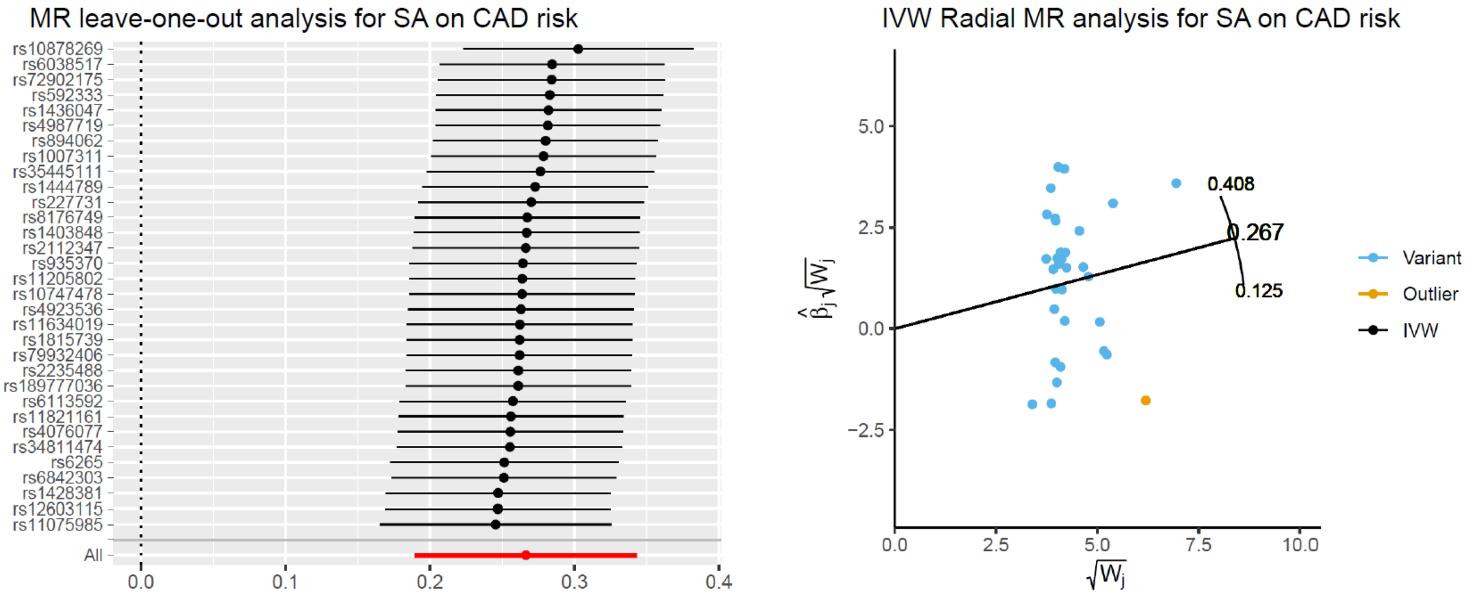


Leave-one-out analysis plots show the Mendelian randomization causal estimates by sequentially dropping one variant at a time, with asymmetry in the plot indicating potential outliers. In the radial plots, the regression line (black line) shows the radial causal estimate for inverse weighted regression models. Variants highlighted in orange were flagged as outliers.

**Supplementary Figure S5. Leave-one-out analysis and radial plot for the effect of coronary artery disease (CAD) on the risk of sleep apnea (SA) using the fixed-effects inverse-variance weighted approach.**

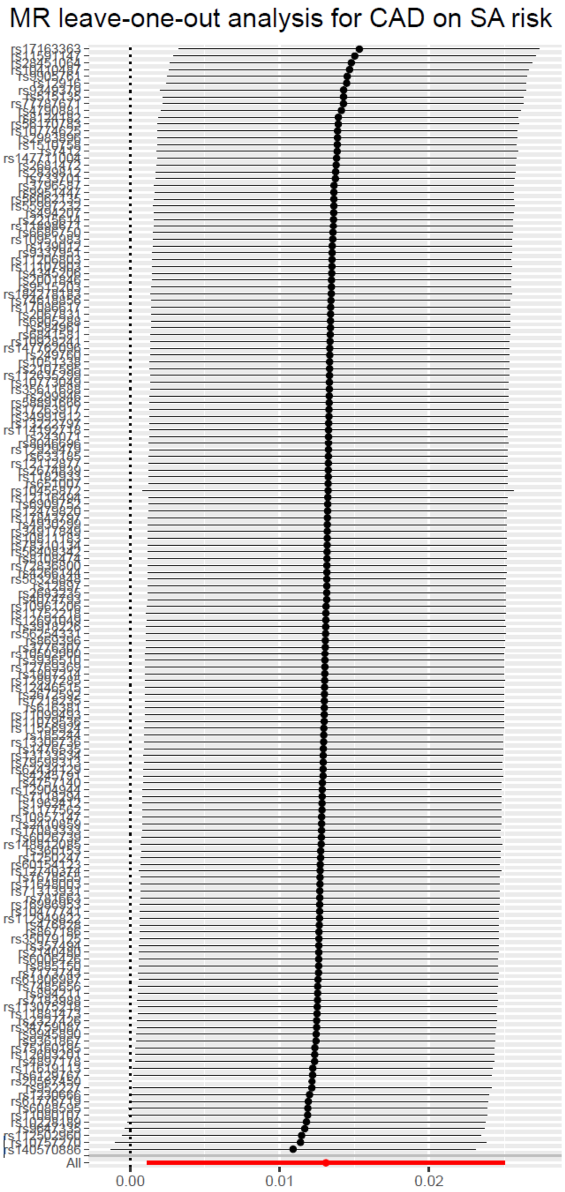


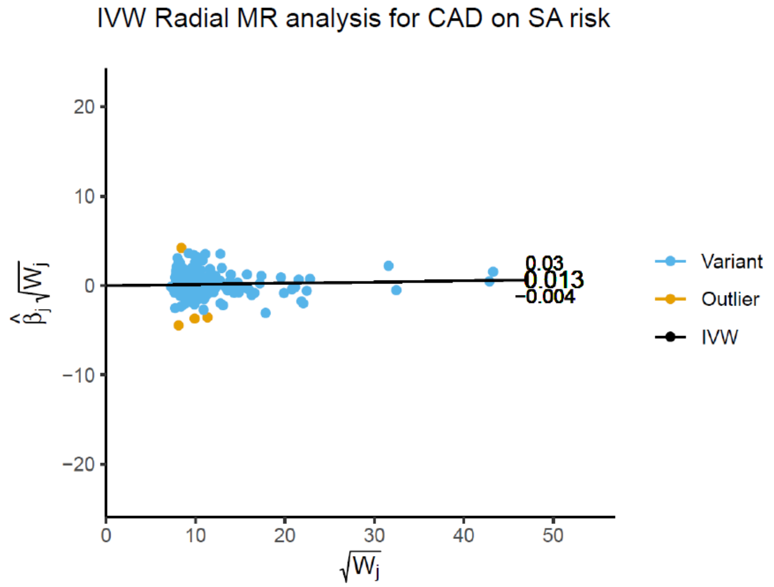


Leave-one-out analysis plots show the Mendelian randomization causal estimates by sequentially dropping one variant at a time, with asymmetry in the plot indicating potential outliers. In the radial plots, the regression line (black line) shows the radial causal estimate for inverse weighted regression models. Variants highlighted in orange were flagged as outliers.

**Supplementary Figure S6. Leave-one-out analysis for the relationships between sleep apnea (SA), Alzheimer's disease (AD) and stroke using the MR-Egger approach.**

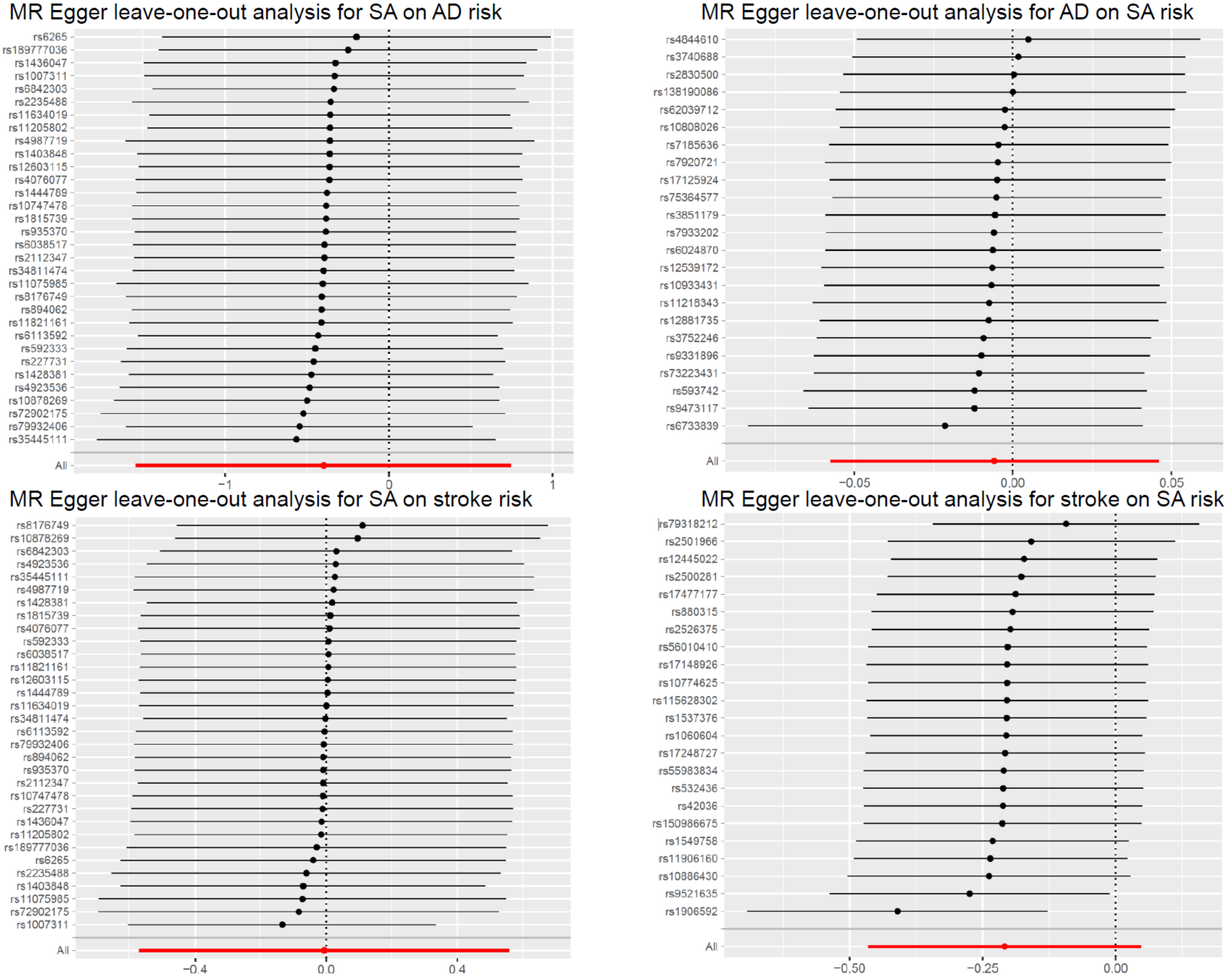


Leave-one-out analysis plots show the Mendelian randomization causal estimates by sequentially dropping one variant at a time, with asymmetry in the plot indicating potential outliers.

**Supplementary Figure S7. Leave-one-out analysis plot for the effect of coronary artery disease (CAD) on the risk of sleep apnea (SA) using the MR-Egger approach.**

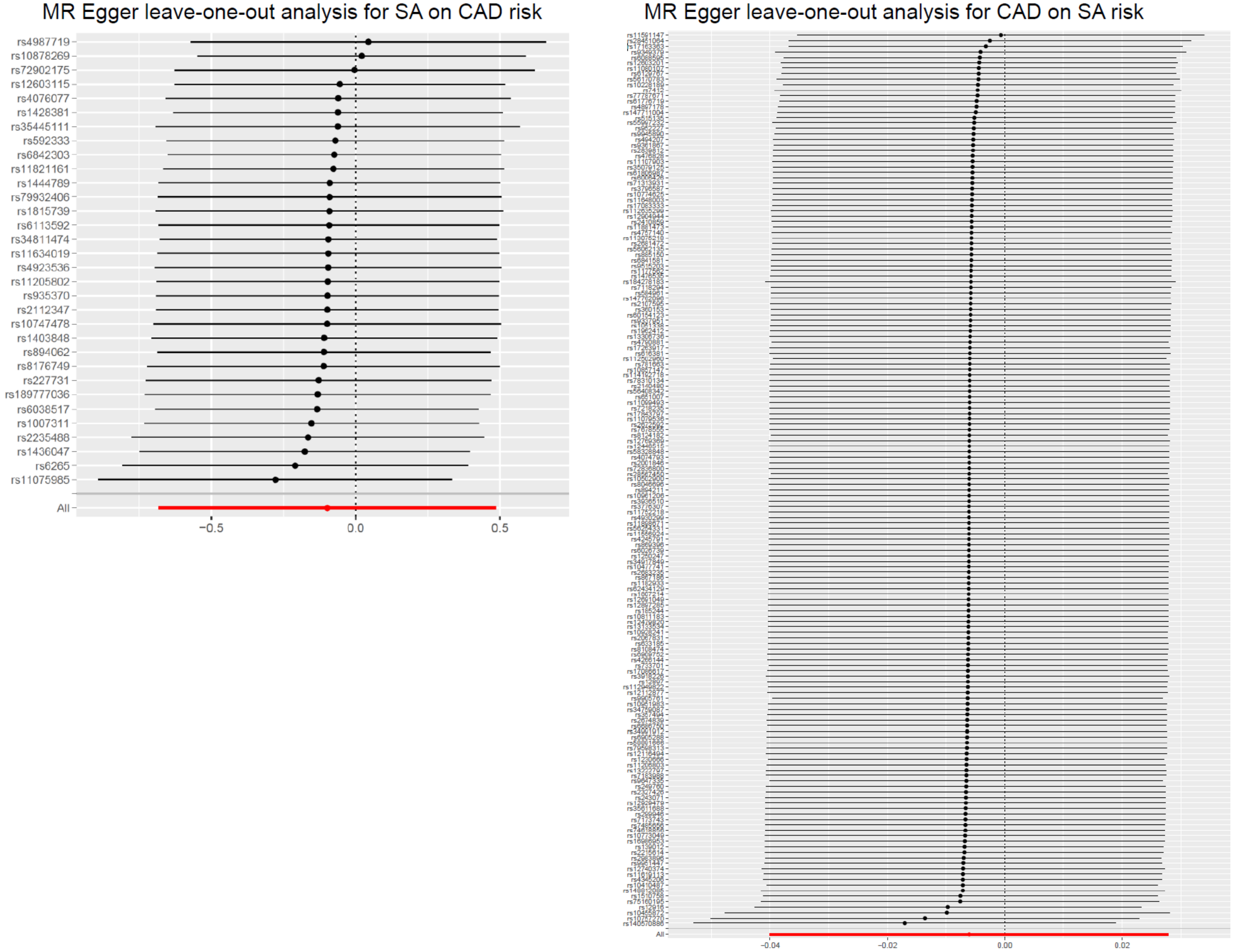


Leave-one-out analysis plots show the Mendelian randomization causal estimates by sequentially dropping one variant at a time, with asymmetry in the plot indicating potential outliers.
